# The 14-protein lung cancer signature: questions of prediction, specificity, and causality

**DOI:** 10.64898/2026.09.08.26362484

**Authors:** Amit Sud, Richard Houlston

## Abstract

Plasma protein signatures may predict future disease but prediction does not necessarily imply disease-specific biology or causation. Distinguishing these possibilities is essential when a circulating signature is proposed as a marker of a pre-malignant state or as a basis for preventive treatment. Pandya *et al.* propose that a 14-protein plasma signature predicts incident lung cancer, identifies individuals who benefit from interleukin-1β blockade for lung cancer prevention, and reflects a causal, interceptable pre-malignant state. We show that two of these claims are more parsimoniously explained by cumulative smoking exposure than by lung-cancer-specific biology. The signature’s incremental discrimination over established clinical risk factors is small, and its associations extend across a broad range of smoking-related and cardiovascular conditions rather than concentrating in the lung. Consistent with this, two-sample Mendelian randomisation provides no evidence that any of the 14 proteins causally influence lung cancer, whereas smoking causally raises circulating levels of seven of them. The claim of predictive therapeutic utility is similarly unsupported. A re-analysis of the CANTOS trial shows no significant treatment-by-signature interaction, providing no evidence that the signature identifies patients who benefit differentially from canakinumab. Taken together, the available human data establish neither a lung-specific, causal pre-malignant mechanism nor a validated basis for treatment selection.

## INTRODUCTION

For a novel biomarker to have clinical value as a predictor, it need not be causal or disease-specific. However, it should be sufficiently strongly associated with outcome to discriminate between affected and unaffected individuals, and it should provide information beyond that already available from routinely measured risk factors, such as age and smoking history. To demonstrate that a biomarker predicts differential treatment benefit, there must be evidence that the relative risk reduction conferred by treatment differs between biomarker-positive and biomarker-negative individuals, formally assessed using a test for treatment-by-biomarker interaction. Finally, to support a causal and potentially interceptable mechanism, the biomarker should lie on the causal pathway to disease rather than represent a downstream consequence of occult disease or a correlate of shared upstream causes such as smoking.

Pandya *et al.* propose that a 14-protein plasma panel, derived from a machine learning model using 2,923 proteins measured in 300 incident lung cancer cases (among 36,074 UK Biobank participants), may be useful both for predicting incident lung cancer and for identifying individuals likely to benefit from interleukin-1β blockade for lung cancer prevention^1^. Here we use additional statistical, epidemiological, and genetic analyses to examine the signature’s predictive value, its basis for treatment selection, and its proposed causal role.

## RESULTS

### Discrimination of incident lung cancer risk

In the held-out set of 75 incident lung cancer cases among 12,025 individuals, the proteomic signature discriminated incident lung cancer to a similar extent as routinely available clinical characteristics (Pandya *et al.* Fig. S2A). Likewise, the combined proteomic and clinical risk factor model did not significantly outperform the established LLPv3 risk model (Pandya *et al.* Fig. 1C). Translating these ROC-AUC estimates into screening-relevant metrics using an equal variance binormal approximation, the detection rate at a 5% false-positive rate (DR5) increases from approximately 34% for LLPv3 to 47% for the combined model. Assuming a 1% disease prevalence (odds 1:99), this corresponds to a modest improvement in post-test odds from approximately 1:15 to 1:11. Overall, the incremental gain over existing risk-stratification tools is small, and its clinical value over current screening approaches remains to be demonstrated.

**Figure 1.**
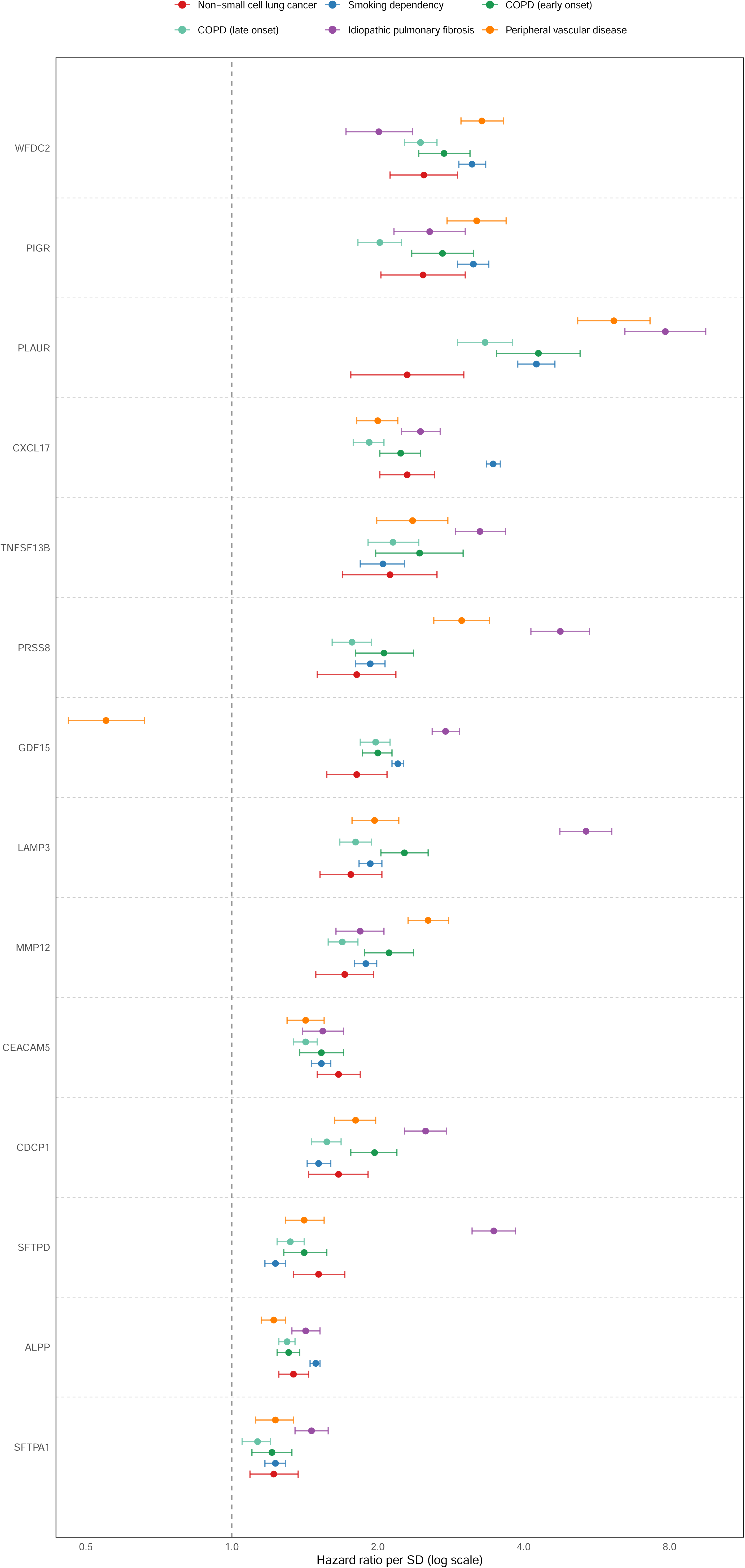
Fourteen-protein signature associations with incident lung, respiratory, vascular and smoking-related disease in UK Biobank. Hazard ratios (HR) per standard deviation (with 95% confidence intervals) for each of the 14 proteins comprising the Pandya *et al*. signature, across five incident-disease endpoints in the UK Biobank Proteome–Phenome Atlas^8^: non-small-cell lung cancer (NSCLC, red), smoking dependency (blue), early-onset chronic obstructive pulmonary disease (green), late-onset chronic obstructive pulmonary disease (lime green), idiopathic pulmonary fibrosis (purple), and peripheral vascular disease (orange). Estimates are from the same cohort and proteomic platform (Olink) as Pandya *et al.*, from Cox models adjusted for age, sex, ethnicity, Townsend deprivation index, body-mass index, smoking status, fasting time, season and blood sample age. Proteins are ordered by their HR for NSCLC.

This modest incremental screening performance beyond readily ascertained clinical factors suggests this protein signature primarily indexes smoking exposure, rather than providing independent predictive information for lung cancer risk. This interpretation is supported by the findings of Xiao *et al.*, who analyzed the same UK Biobank cohort using the same Olink platform^2^. Notably, the two most influential proteins in Xiao’s smoking model, ALPP and CXCL17, are both included in the cancer panel reported by Pandya *et al.* Each of these proteins discriminates current from never-smokers with high accuracy (AUC 0.88 and 0.87, corresponding to DR5 values of 51% and 48% respectively). Thus, either protein alone distinguished smokers from never-smokers more effectively than the full 14-protein signature distinguished future lung cancer cases from controls.

More broadly, six of the 14 proteins in the Pandya *et al.* signature are also components of Xiao *et al*.’s proteomic Smoking INdex (pSIN), for which smoking history accounts for 66% of the variance. pSIN is associated with incident lung cancer (HR 1.97 per 1 SD, 95% CI 1.83–2.11; DR5 = 17%) but is also associated almost as strongly with chronic obstructive pulmonary disease (COPD) (HR 1.72 per 1 SD, 95% CI 1.67–1.78; DR5 = 14%) as well as other smoking-related conditions including peripheral artery disease (PVD), ischaemic heart disease (IHD) and all-cause mortality. This pattern suggests that pSIN captures the broader systemic consequences of smoking rather than lung cancer specifically. Taken together, these observations indicate that the 14-protein signature does predict incident lung cancer, but adds only modestly to clinical risk factors. Moreover, the incremental information it provides may reflect a more accurate smoking exposure, rather than biology specific to lung carcinogenesis.

### Prediction of response to interleukin-1β blockade for lung cancer prevention

For a biomarker to demonstrate predictive therapeutic utility, the magnitude of treatment response must differ significantly according to biomarker status, as assessed by a formal treatment-by-marker interaction test. In their post-hoc analysis of the CANTOS trial, Pandya *et al*. reported an odds ratio for lung cancer following canakinumab treatment of 0.52 (95% CI 0.31–0.86) in the high-signature group and 0.91 (95% CI 0.34–2.48) in the low-signature group. Using the method of Altman and Bland to compare two independent estimates on the log scale^3^, the ratio of odds ratios is 0.57 (95% CI 0.19 to 1.75), corresponding to a treatment-by-signature interaction of z = −0.98 (*P* = 0.33). Thus, there is no statistically significant evidence that the effect of canakinumab differs between the two signature groups. Although the authors acknowledge that the interaction analysis is underpowered, this limitation also means that the available data do not establish that the signature identifies individuals who derive greater benefit from IL-1β blockade. Consequently, the proposed prevention strategy cannot be justified on the basis of differential treatment benefit between subgroups.

### Causal interpretation of the human data

The paper goes beyond risk prediction to propose a specific causal mechanism: that the 14 proteins reflect a tumour-promoting alveolar niche driven by particulate matter, oncogenic EGFR clones, and IL-1β signalling, representing a pre-malignant state that can be intercepted therapeutically. The accompanying mouse experiments are presented as support of this framework. However, establishing this mechanism in humans requires excluding explanations for the observed protein associations, which the study does not do.

An important alternative explanation is that the circulating proteins reflect occult lung cancers already present at the time of sampling rather than a pre-malignant state. For samples collected one to three years before diagnosis, elevated protein concentrations could arise from subclinical tumour burden, tumour-induced host response, or other consequences of established disease. The external validation cohorts do not distinguish between these possibilities. In the Lung Cancer Cohort Consortium, the mean interval between sampling and diagnosis was only 1.6 years^4^, a time frame within which many participants would be expected to harbour undiagnosed cancers. The issue is even more pronounced in

TALENT, where the median lead time was only 144 days and 87.6% of cases were stage IA at diagnoisis^5^. Consistent with an imminent-disease signal, the proteomic model provides additional discrimination over established screening models only shortly before diagnosis (Pandya *et al*., Fig. 1D). The unreported but decisive test, that would distinguish a pre-malignant biomarker from an occult-disease biomarker, is whether predictive performance is maintained in individuals sampled years before diagnosis.

The authors’ own longitudinal analyses also favour this interpretation. In the full UKCTOCS cohort, CXCL17 and WFDC2 are elevated in cases up to five years before diagnosis (Pandya *et al.*, Fig. 1E), whereas CEACAM5 rises predominantly in the final two years before diagnosis. However, in never-smokers, none of these proteins differs between cases until approximately two years before diagnosis (Pandya *et al.*, Fig. S2B). The disappearance of the apparent long-term signal after restricting the analysis to never-smokers suggests that the early separation in the full cohort is largely driven by smoking-related differences rather than by a shared pre-malignant biological process. The remaining late signal is dominated by CEACAM5, an established tumour-derived serum marker and, in tissue, a marker of the invasive lung adenocarcinoma, the predominant subtype in never-smokers and women, consistent with the UKCTOCS cohort^6,7^. Collectively, these findings are more readily explained by a combination of smoking-related inflammation at longer lead times and tumour-derived protein release at diagnosis rather than by the existence of a pre-malignant proteomic state.

A second alternative explanation is that the signature reflects systemic smoking-related damage rather than a biological process causally upstream of lung cancer. In the UK Biobank Proteome–Phenome Atlas (the same cohort and platform as Pandya *et al*.; incident-disease hazard ratios adjusted for smoking status and other risk factors), the 14 proteins are not selectively associated with lung cancer^8^. Instead each ranks the strongest protein-level predictors for a broad range of smoking-related conditions, including smoking dependency (**Fig. 1**), and collectively the proteins are markedly enriched among the strongest predictors of cardiovascular disease (CVD), including IHD, heart failure, stroke and PVD (**Fig. 2**). Associations of this breadth, despite adjustment for smoking, are difficult to reconcile with a lung-specific tumour-promoting niche (which would be expected to concentrate its strongest associations in pulmonary rather than CVD) and instead suggest that the signature captures systemic smoking-related injury that is correlated with lung cancer through shared causation. Such a marker would be expected to improve on questionnaire-based risk factors, because circulating proteins provide a more faithful measure of cumulative biological exposure than self-reported smoking history. Six of the 14 proteins are components of Xiao *et al.*’s protein Smoking Index (pSIN), a reversible marker of cumulative exposure that declines gradually after smoking cessation while predicting lung cancer and multiple smoking-related diseases (Xiao *et al*., Fig. 5)^2^. Even after adjustment for pack-years and time since cessation, pSIN remains strongly associated with disease risk among former smokers. Individuals whose proteomic profiles have returned to never-smoker levels have substantially lower risks of lung cancer (HR = 0.37), COPD (HR = 0.33), PVD (HR = 0.54) and all-cause mortality (HR = 0.52). Thus, improved prediction beyond conventional clinical risk factors does not itself imply lung cancer specific biology. This interpretation is reinforced by Tsuo *et al*.’s independent analysis of the same UK Biobank Pharma Proteomics Project (UKB-PPP) cohort^9^. All 178 proteins associated with lung cancer were also independently associated with smoking status, and adjustment for smoking reduced the number of significantly associated proteins from 324 to 33, the largest attenuation of any disease examined. The authors additionally derived a proteomic smoking score, conceptually analogous to Xiao *et al*.’s pSIN^2^, which failed to discriminate lung cancer cases from controls among never-smokers or improve their prediction over baseline. Tsuo *et al*. conclude that such smoking-associated proteins are best interpreted as environmental sensors reflecting cumulative smoking exposure, rather than causal drivers of disease.

**Figure 2.**
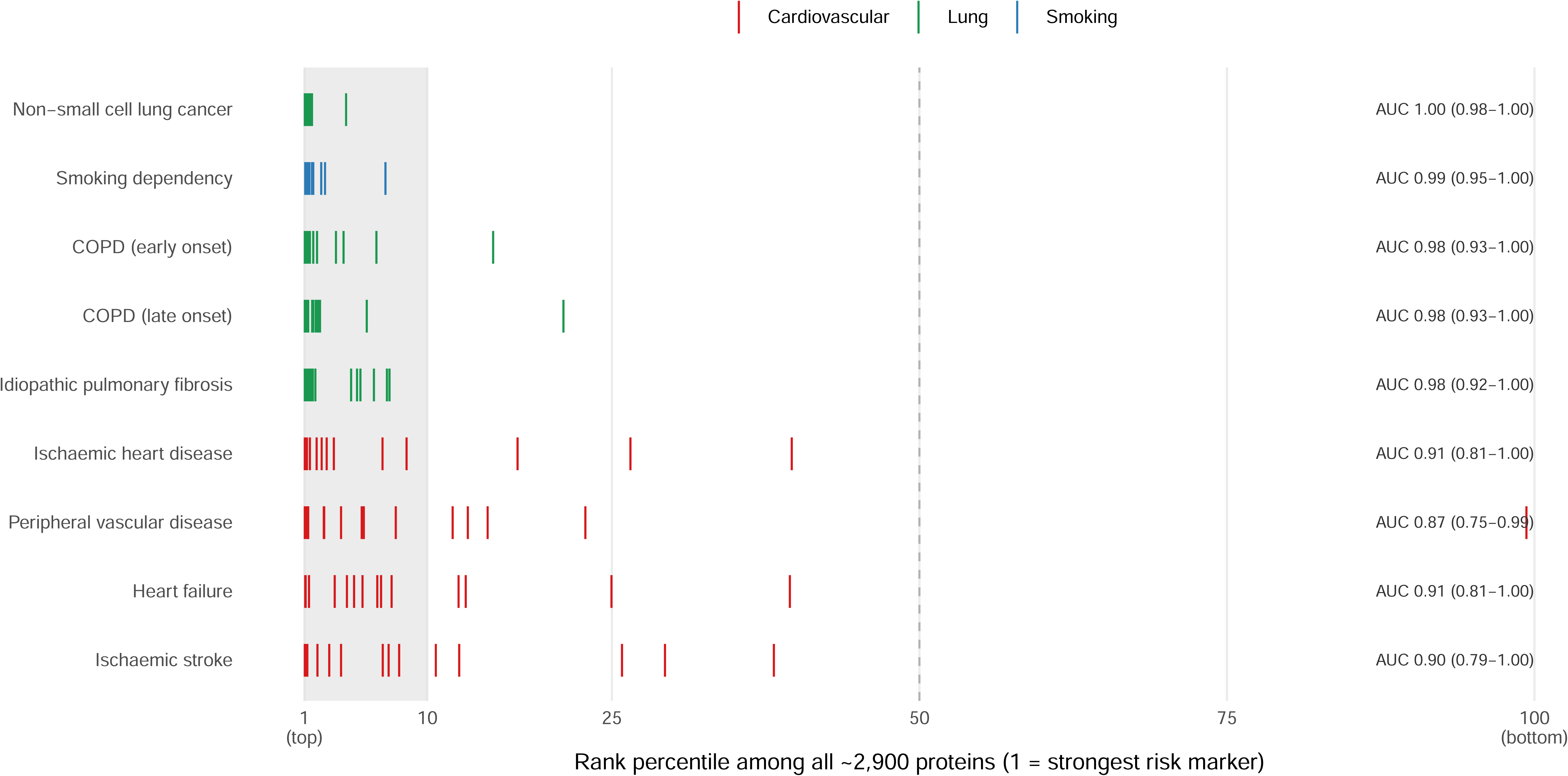
Rank of the 14 signature proteins among protein-level risk markers of lung, smoking-related and cardiovascular disease in UK Biobank. For each disease, all ∼2,900 plasma proteins measured in the UK Biobank Proteome–Phenome Atlas were ranked by the strength of their association with incident disease (z = log hazard ratio / standard error, from Cox models adjusted for age, sex, ethnicity, Townsend deprivation index, body-mass index, smoking status, fasting time, season and blood sample age)^8^. Each vertical tick marks the rank position of one of the 14 signature proteins; the left edge is the strongest risk marker and the shaded band the top 10%. Diseases are coloured by group (lung, green; smoking dependency, blue; cardiovascular, red) and ordered by the median rank of the 14 proteins. For each disease, AUC is the probability that a randomly chosen signature protein outranks a randomly chosen non-signature protein (Mann–Whitney; Hanley and McNeil 95% confidence interval).

Pandya *et al*. note that the signature does not decline following surgical resection in the TRACERx cohort and interpret this as evidence against a tumour-derived origin. However, this finding is equally compatible with a smoking-related signature. Surgical resection removes the tumour but not the cumulative biological consequences of smoking, and proteomic smoking indices such as pSIN decline slowly with more than 80% of former smokers remaining above the smoking threshold even ten years after cessation^2^. Even if framed as field cancerisation, in which persistent molecular alteration occurs across smoke-exposed epithelium, persistence alone would reflect cumulative exposure rather than establish that the 14 proteins causally mediate tumour promotion; that distinction requires causal analysis, not plasma persistence alone. Consistent with a smoking-exposure interpretation, the signature discriminates only weakly between cases and controls in TALENT, a cohort comprising 93% never-smokers and only 7% former smokers (Pandya *et al.*, Fig. S2F)^5^. Once smoking exposure is largely absent, much of the proteomic signal also disappears. Distinguishing a causal mediator from a correlated marker requires analyses that directly address this alternative explanation. One approach would be to test whether the association between the 14-protein signature and lung cancer remains after adjustment for an established proteomic smoking index such as pSIN, an analysis that appears feasible within the available cohort but is not reported. Likewise, the authors do not report the associations of their signature with smoking status or with a broad spectrum of pulmonary and non-pulmonary diseases and mortality that our analyses suggest. Providing these analyses would substantially clarify whether the signature captures lung cancer specific biology or more general smoking related injury.

The accompanying mouse experiments provide a coherent mechanistic hypothesis and are of genuine scientific interest. However, the human observational data remain consistent with two simpler explanations: that the proteins reflect occult lung cancer or that they capture systemic smoking-related damage. To formally test the authors’ central causal claim, we performed two-sample Mendelian randomisation for all 14 panel proteins, using *cis*-protein quantitative trait loci from the same UKB-PPP resource as genetic instruments. Outcome data were obtained from the International Lung Cancer Consortium (ILCCO) for lung cancer (29,266 cases and 56,450 controls) and the GWAS & Sequencing Consortium of Alcohol and Nicotine use for smoking behaviour (249,171 individuals)^10,11^. No protein showed statistically significant evidence of a causal effect on lung cancer or smoking behaviour (**Figure 3**, **Supplementary Table 1**, **Supplementary Table 2**)). However, genetic instruments for smoking initiation and cigarettes per day showed causal effects on plasma levels of seven of the 14 panel proteins (**Figure 3** and **Supplementary Table 3**). This asymmetry, smoking causally shaping proteins, but not vice versa is the pattern expected if the panel indexes smoking exposure rather than mediating its consequences. Together, these findings are consistent with smoking-related confounding, rather than a causal contribution of the proteins to malignancy, and provide no support for the proposed causal, pre-malignant mechanism in humans. Ascertaining whether the signature predicts cancer specifically in never-smokers with long intervals between sampling and diagnosis would help establish whether occult disease also contributes, and is of particular importance given the rising incidence of lung cancer in never-smokers.

**Figure 3.**
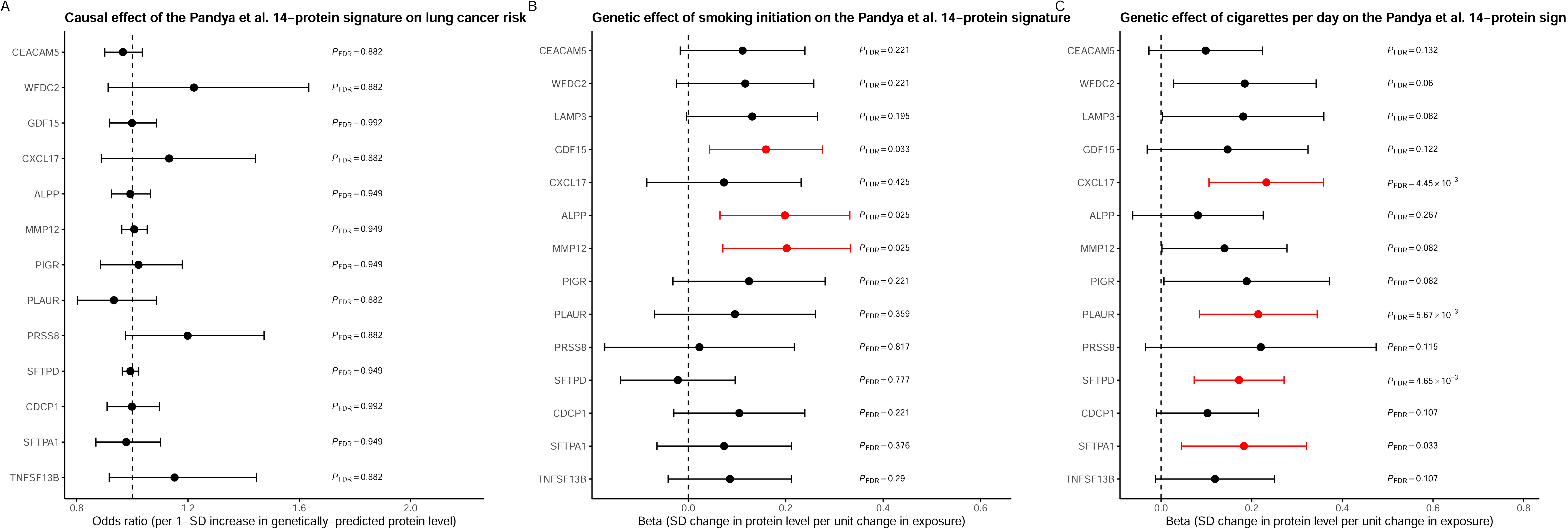
Mendelian randomisation evidence for causal relationships between the Pandya et al. 14-protein signature, smoking behaviour, and lung cancer. (A) Causal effect of each of the 14 panel proteins on lung cancer risk (odds ratio, 95% CI), using *cis*-protein quantitative trait loci as genetic instruments (Supplementary Table 1). (B) Causal effect of smoking initiation and (C) cigarettes per day on plasma levels of the Pandya et al. 14-protein signature (beta, 95% CI), using genetic instruments for each smoking phenotype (Supplementary Table 3). Beta represents SD change in protein level per unit increase in the respective exposure. Red indicates FDR-significant estimates within each panel (Benjamini-Hochberg, applied across all tested lung cancer outcomes for panel A, and separately within each smoking phenotype for panels B–C); black indicates non-significant. FDR-adjusted P values are shown alongside each estimate. Dashed lines denote the null (odds ratio=1 for panel A; beta=0 for panels B–C).

## DISCUSSION

Pandya *et al*. have made an important contribution by integrating large-scale population proteomics with mechanistic models of carcinogenesis. The 14-protein signature may ultimately prove valuable as a tool for early detection of subclinical lung cancer, and despite the failure of canakinumab in therapeutic trials of metastatic and adjuvant NSCLC^1213^, IL-1β inhibition may have a role in cancer prevention. However, the improvement in prediction over established clinical risk factors is small at best, the evidence for treatment effect modulation does not satisfy the statistical criterion of a significant biomarker-by-treatment interaction, and the causal interpretation of the human data is better explained by smoking-related confounding than by a causal, pre-malignant mechanism. Addressing these points would allow readers to assess more clearly what the signature does and does not establish.

## METHODS

### Detection rate and post-test probability conversion

Reported AUCs were converted to detection rate at a fixed 5% false-positive rate (DR5) under the equal-variance binormal ROC model. Post-test odds and positive predictive value at a specified disease prevalence were derived as (DR/FPR) × prevalence/(1−prevalence).

### Treatment-by-biomarker interaction test

Canakinumab’s treatment effect between biomarker-signature-high and -low subgroups was assessed using the method of Altman and Bland. Each subgroup’s log(OR) standard error was calculated from its reported 95% CI, and the two log(OR) estimates were compared via a z-test on their difference, with variance equal to the sum of the two subgroup variances.

### Per-protein multi-trait association profiling

Smoking-adjusted incident hazard ratios (per SD) for each of the 14 signature proteins were obtained from the UKB-PPP for six endpoints: non-small cell lung cancer, smoking dependency, early- and late-onset COPD, idiopathic pulmonary fibrosis, and peripheral vascular disease [adjusted for age, sex, ethnicity, Townsend deprivation index, BMI, smoking status, fasting time, season, and blood sample age].

### Enrichment analysis

To assess the representation of the 14 protein signature among genome-wide disease-associated proteins, all ∼2,900 proteins in the UKB-PPP Atlas were ranked by association strength (z = log(HR)/SE) for each of nine endpoints spanning lung, smoking, and CVD, and the percentile rank of each signature protein within this distribution was determined. Enrichment was quantified as the area under the curve (AUC) of a Mann-Whitney U statistic comparing signature versus non-signature protein ranks, with Hanley-McNeil 95% confidence intervals (approximate, as protein measurements are not independent).

### Mendelian randomisation

We performed two-sample Mendelian randomization (MR) to assess whether the 14 proteins in the Pandya et al. panel have causal effects on lung cancer risk and smoking-related behaviours, and, in the reverse direction, whether smoking behaviours have causal effects on levels of these proteins. *Cis*-protein quantitative trait loci were obtained from the UKB-PPP discovery cohort (n=34,557, European ancestry). For each protein, genome-wide significant variants (*P*<5×10⁻⁸) within 1Mb of the transcriptional start site were retained, excluding those with minor allele frequency ≤0.01, INFO score ≤0.8, or F-statistic ≤10. Independent instruments were selected by LD clumping (r²≤0.001) using the European-ancestry UK Biobank LD reference panel (n=337,545).

Outcome data comprised seven phenotypes. Lung cancer data were obtained from the ILCC, which excludes UK Biobank participants: lung cancer overall, adenocarcinoma, squamous cell carcinoma, and lung cancer in ever- and never-smokers (GWAS Catalog accessions GCST004748, GCST004744, GCST004750, GCST004749, and GCST004747, respectively)^10^. Smoking behaviour data were obtained from GSCAN GWAS summary statistics excluding UK Biobank and 23andMe participants: smoking initiation and cigarettes per day^11^. For the reverse direction, genome-wide significant variants for these same two phenotypes (10 and 8 independent loci respectively, were against all 14 proteins^14^.

Exposure and outcome data were harmonized using the TwoSampleMR R package, aligning effect alleles and resolving palindromic variants by allele frequency where unambiguous^15^. Variants with stronger evidence of association with the outcome than the exposure were excluded using TwoSampleMR’s Steiger filtering^16^. Causal estimates were derived using the Wald ratio (single-instrument proteins) or inverse-variance-weighted method (≥2 instruments), using the MendelianRandomization R package. Where ≥3 instruments were available, weighted median, weighted mode, and MR-Egger were additionally applied to assess robustness to invalid instruments and horizontal pleiotropy, and Radial MR was used to test each instrument’s individual contribution to heterogeneity via a Bonferroni-corrected test, with outlier-excluded sensitivity estimates reported where an outlier was detected^17^. Estimates for binary outcomes are reported as odds ratios; estimates for continuous outcomes (cigarettes per day, and protein levels in the reverse direction) are reported as beta coefficients (SD units) and are not exponentiated. *P*-values were corrected for multiple testing using the Benjamini-Hochberg false discovery rate method.

## Data Availability

All data produced in the present work are contained in the manuscript

https://proteome-phenome-atlas.com/

https://broad-alkesgroup-ukbb-ld.s3.amazonaws.com/UKBB_LD/

https://genome.psych.umn.edu/research/gscan

https://www.ebi.ac.uk/gwas/

## ACKNOWLEDGEMENTS

A.S. is in receipt of a Wellcome Trust Early Career Award (227000/Z/23/Z). R.H. acknowledges grant support from Cancer Research UK (C1298/A8362) and the Wellcome Trust (214388).

## AUTHOR CONTRIBUTIONS

A.S. and R.H. researched, reviewed, drafted and edited the manuscript.

## CODE AVAILABILITY

Code to perform statistical analyses and figure generation in this manuscript is available at (https://github.com/amitsud/14_protein_signature_lung_cancer)

## DATA AVAILABILITY

All data produced in the present work are contained in the manuscript. The protein–disease association data used in this analysis are openly available from the UK Biobank plasma proteome atlas via the interactive web portal at https://proteome-phenome-atlas.com/^8^. The underlying UK Biobank resource is available to approved researchers via application at https://www.ukbiobank.ac.uk/. *Cis*-protein quantitative trait loci for the Mendelian randomisation analyses were obtained from the UK Biobank Pharma Proteomics Project discovery cohort summary statistics^14^. Lung cancer outcome data are available from the GWAS Catalog (https://www.ebi.ac.uk/gwas/, accessions GCST004748, GCST004744, GCST004750, GCST004749, and GCST004747)^10^. Smoking behaviour summary statistics, excluding UK Biobank and 23andMe participants, are available from the GWAS & Sequencing Consortium of Alcohol and Nicotine Use via the University of Minnesota Digital Conservancy (https://genome.psych.umn.edu/research/gscan)^11^. The linkage disequilibrium reference panel used for instrument clumping is available from the Pan-UK Biobank resource (https://broad-alkesgroup-ukbb-ld.s3.amazonaws.com/UKBB_LD/).

